# Influence of age on the effectiveness and duration of protection in Vaxzevria and CoronaVac vaccines

**DOI:** 10.1101/2021.08.21.21261501

**Authors:** Thiago Cerqueira-Silva, Vinicius de Araújo Oliveira, Julia Pescarini, Juracy Bertoldo Júnior, Tales Mota Machado, Renzo Flores-Ortiz, Gerson Penna, Maria Yury Ichihara, Jacson Venâncio de Barros, Viviane S. Boaventura, Mauricio L. Barreto, Guilherme Loureiro Werneck, Manoel Barral-Netto

## Abstract

**Background:** High rates of virus transmission and the presence of variants of concern can affect vaccine effectiveness (VE). Both conditions occur in low-income countries, which primarily use viral vector or inactivated virus vaccine technologies. Such countries conducted few VE analyses, and most lack the power to evaluate effectiveness in subgroups.

**Methods:** The present retrospective cohort study evaluated the effectiveness of Vaxzevria and CoronaVac vaccines for COVID-19-related infection in 75,919,840 Brazilian vaccinees from January 18 to July 24, 2021.

Study outcomes included documented infection with severe acute respiratory syndrome coronavirus 2 (SARS-CoV-2), COVID-19–related hospitalization, ICU admission, and death. We estimated VE using Cox regression adjusted for individual demographic characteristics.

**Results:** Vaccination with Vaxzevria or CoronaVac was effective against SARS-CoV-2 infection and highly effective against hospitalization, ICU admission, and death in individuals up to 79 years. From 80-89 years of age, Vaxzevria led to 89.9%(95CI:87.7-91.7) VE against death versus 67.2%(95CI:63.6-70.5) for CoronaVac. Above 90 years, 65.4%(95CI:46.1-77.8) protection was conferred to Vaxzevria-vaccinated individuals versus 33.6%(95CI:21.9-43.5) in CoronaVac-vaccinated individuals. Furthermore, the post-vaccination daily incidence rate shows a stepwise increase from younger to elder decades of life.

**Conclusions:** Vaxzevria demonstrated overall effectiveness against severe COVID-19 up to 89 years and CoronaVac up to 79 years of age. There is a stepwise effectiveness reduction for both vaccines for each decade of life. Our results suggest that individuals aged 80 years or older may benefit from an expedited booster dose. Ongoing evaluations, including any additional vaccines authorized, are crucial to monitoring long-term vaccine effectiveness.

## Background

Several COVID-19 vaccines have proved efficacious, and many of them are in extensive use around the world.^1–4^ While high-income countries preferentially administer mRNA-based vaccines, lower- and middle-income countries have employed vaccines based on viral vectors or inactivated virus technologies. A timely evaluation of the effectiveness of the currently available vaccines across different regions is essential for a comprehensive understanding of vaccine impact, considering significant variations in vaccination schedules, virus transmission, and the emergence of viral variants, in addition to social and cultural standards and local health system conditions.

Brazil is one of the countries most affected by the pandemic, with high rates of transmission. The Brazilian COVID-19 vaccination program initially relied on Vaxzevria/Fiocruz (previously Oxford-AstraZeneca or ChAdOx-1), approved in 181 countries, and Sinovac’s CoronaVac/Butantan, approved in 39 countries.^5^ The recommended inter doses interval in Brazil for Vaxzevria is 12 weeks versus 2-4 weeks for CoronaVac.^6^ The period between doses of Vaxzevria has varied in several countries.^7^ CoronaVac has been applied at distinct intervals,^1,8^ making direct comparisons difficult. Additionally, several early publications on vaccine effectiveness (VE) evaluated only the initial dose or were limited to analyzing effectiveness against symptomatic infection^9,10^ and hospitalization^10,11^, i.e., ICU admission and death were not addressed.

Nationwide evaluations of the effectiveness of COVID-19 vaccines in Brazil offers advantages, as this country’s large population is distributed throughout several regions with considerable differences in socio-economic aspects and access to medical facilities. Nonetheless, data collection systems are identical throughout the entire country, offering a comprehensive source of data to perform a countrywide VE evaluations. The COVID-19 vaccination campaign was initiated nationwide on January 18, 2021. By July, many vaccinees had received either Vaxzevria/Fiocruz or CoronaVac/Butantan vaccines, allowing for a detailed evaluation of the effectiveness of both vaccines while considering several outcomes and stratified age ranges, making it possible to examine in detail specific age effects previously not investigated.

A significant issue regarding the VE of vaccines against COVID-19 is the degree of circulation of distinct SARS-CoV-2 variants of concern (VOC) in different regions. During the present study, the Gamma variant was the most frequent in all regions of Brazil.^12^ Importantly, the literature contains few reports on the VE of Vaxzevria and Coronavac against the Gamma variant.^1,10,13^

The present study aimed to evaluate the effectiveness of Vaxzevria and Coronavac vaccines in 75,919,840 Brazilian vaccinees with respect to several different outcomes: COVID-19 related infection, hospitalization, ICU admission and death, between January 18 and July 24, 2021.

## Methods

### Study design and datasets

We conducted a retrospective cohort using individual-level information on demographic, clinical characteristics, and SARS-COV-2 laboratory tests from Brazilian datasets. The Brazilian Ministry of Health Department of Informatics provided unidentified datasets of the COVID-19 Vaccination Campaign (SI-PNI), the Acute Respiratory Infection Suspected Cases (e-SUS-Notifica), and the Severe Acute Respiratory Infection/Illness (SIVEP-Gripe). A key-coded individual identification number present in the three datasets was used for a deterministic linkage and then removed from the resulting dataset used in our analyses. No personally identifiable data was accessed at any stage. Codebooks, scripts and public dataset version will be available at https://vigivac.fiocruz.br SI-PNI is a data warehouse with all the vaccine doses administered by health services in Brazil. From SI-PNI, we extracted information on the type and date of COVID-19 vaccine administration. Brazilian population estimates for 2021 corrected by the all-cause deaths reported in 2020 were retrieved from previous study.^14^

The e-SUS-Notifica is a national online health surveillance information system for register of acute respiratory infections and suspected/confirmed COVID-19 and has been used as a data source for epidemiological research.^15^

SIVEP-Gripe is the national system for register of SARI-related hospitalizations and deaths that was created during the H1N1 pandemic in 2009 and have been widely used as a source for epidemiological studies.^16–18^ All COVID-19 related SARI hospitalizations and deaths (independent of hospitalization) are registered in the system. Open versions of all datasets are available <u>opendatasus</u>.

From both SIVEP-Gripe and e-SUS-Notifica, we extracted information on the date of symptom onset, RT-PCR and antigen test results for SARS-CoV-2. From SIVEP-Gripe, we obtained data of hospitalization, admission to ICU, and hospitalization outcome (discharge or death).

### Study population

We included all individuals who received the COVID-19 vaccine first dose between January 18, 2021, and July 24, 2021. They were followed retrospectively to assess infection, hospitalization, admission to ICU, and death with a laboratory-confirmed diagnosis of SARS-CoV-2 up to July 24, 2021.

We excluded individuals (i) vaccinated with vaccines besides Vaxzevria or CoronaVac, (ii) with inconsistent vaccine records (i.e., individuals who received the second dose without the first dose, doses from different vaccines or interval between doses less than 14 days), (iii) with confirmed COVID-19 before the date of vaccine administration, and (iv) with missing data for essential covariates (sex or age).

### Exposure and outcomes

We defined vaccination status for each vaccine based on the time elapsed since the administration of a vaccine dose:

1. ≤13 days after the first dose (the reference period)
2. ≥14 days after the first dose and without the second dose (partially vaccinated)
3. ≥14 days after the second dose (fully vaccinated)

The reference period was defined based on results of a Phase III randomized controlled trial^8^ and three test-negative case-control studies.^11,19,20^ The time-lapsed between the date of the first dose and the development of an effective immune response is used to detect bias in test-negative case-control studies to estimate vaccine effectiveness, the theoretical frame for such use has been discussed by Hitchings et al.^21^ We also analyzed vaccine effectiveness for 1 to 13 days after the second dose, with the results presented in supplementary table S2.

Laboratory confirmation of COVID-19 with a positive RT-PCR or antigen test result was required for inclusion in the analyses. The outcomes analyzed were infection, hospitalization, admission to an intensive care unit (ICU), and death by COVID-19. For all outcomes we considered the time between the first or second vaccination date up to the symptom’s onset. Death was considered at any time regardless of prior hospitalization. ICU admission was considered at any time point between the admission and the discharge or death dates.

### Statistical analyses

In the primary analysis, we used a Cox regression model to estimate the hazard ratio (HR) of COVID-19 infection, hospitalization, ICU admission, and death for partially and fully vaccinated individuals, adjusted for age, sex, region of residence, socioeconomic status, and month of the 1st dose. We used the Brazilian Deprivation Index (*Índice Brasileiro de Privação*), a municipality-level measure of material deprivation, as an indicator of socioeconomic status.^22^ We estimated vaccine effectiveness (VE) as 1-HR, obtained from a model including all covariates, and reported as a percentage. We also reported the crude VE for each outcome. In addition, we performed a stratified analysis by age groups (<60, 60–69, 70–79, 80–89, ≥90 years).

To evaluate the validity of our assumptions, we compared the hazards of infection hospitalization, ICU admission and death in the reference period for each vaccine and visually assessed the daily number of cases after each dose of vaccine. To assess the robustness of our findings, we repeated the principal analysis defining as the reference period the time elapsed up to 10 days after the date of the first dose, as it is expected that the vaccines ‘protection increases with time. Additionally, we examined the VE for hospitalization, ICU admission and death using clinical suspected cases besides laboratory confirmed ones.

Analyses were performed using the R statistical software and H2O package.^23,24^ Descriptive statistics were presented as frequencies and percentages. We used the 95% confidence intervals (CI) of the estimated measures of association for interpreting the findings.

## RESULTS

From January 18 to July 24, 2021, 96,193,523 individuals received at least one dose of one of the two COVID-19 vaccines analyzed in this study, and 77,652,891 (80.7%) met the initial eligibility criteria. After excluding individuals who had COVID-19 before vaccination, with problems with registries of vaccine doses administration or lack of information on sex, 75,919,840 individuals were selected for the analysis (Figure 1). The majority (66.1%, n=50,167,827 individuals) received at least one dose of Vaxzevria and the remaining (33.9%, n=25,752,013 individuals) received at least one dose of CoronaVac. The majority of our cohort comprised women (54.8%) and individuals aged 60 years or older (36.8%). Compared to individuals that received CoronaVac, individuals that received Vaxzevria were younger (24.2% vs. 61.5% of individuals aged 60 years or older), and a lower proportion had completed the full vaccine schedule (20.2% vs. 78.6%). Vaccination with CoronaVac occurred mainly from January to April 2021, while Vaxzevria was administered predominantly after March 2021 (Figure 2). Among those who received the second dose, the median time between doses was 84 days (IQR 82–90) for Vaxzevria and 27 days (IQR 21– 28) for CoronaVac (Supplementary Table S1).

**Figure 1.**
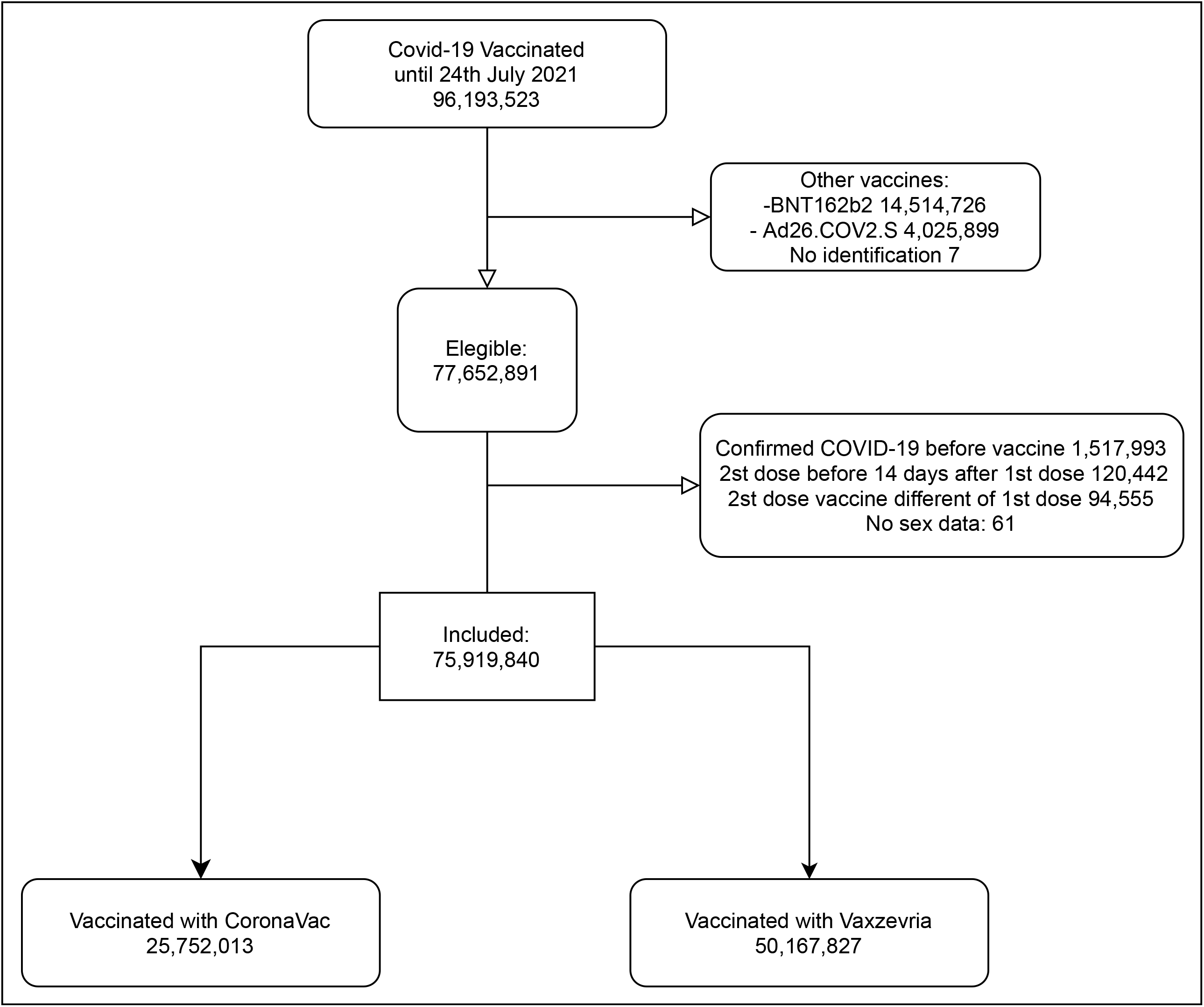
Flowchart of the selection of the study individuals vaccinated between January 18 and July 24, 2021. Eligible participants received at least one dose of CoronaVac or Vaxzevria vaccine between January 18 and July 24, 2021. We excluded persons with confirmed COVID-19 diagnosis in 2021 before the first dose and all persons with different vaccines from CoronaVac or Vaxzevria

**Figure 2.**
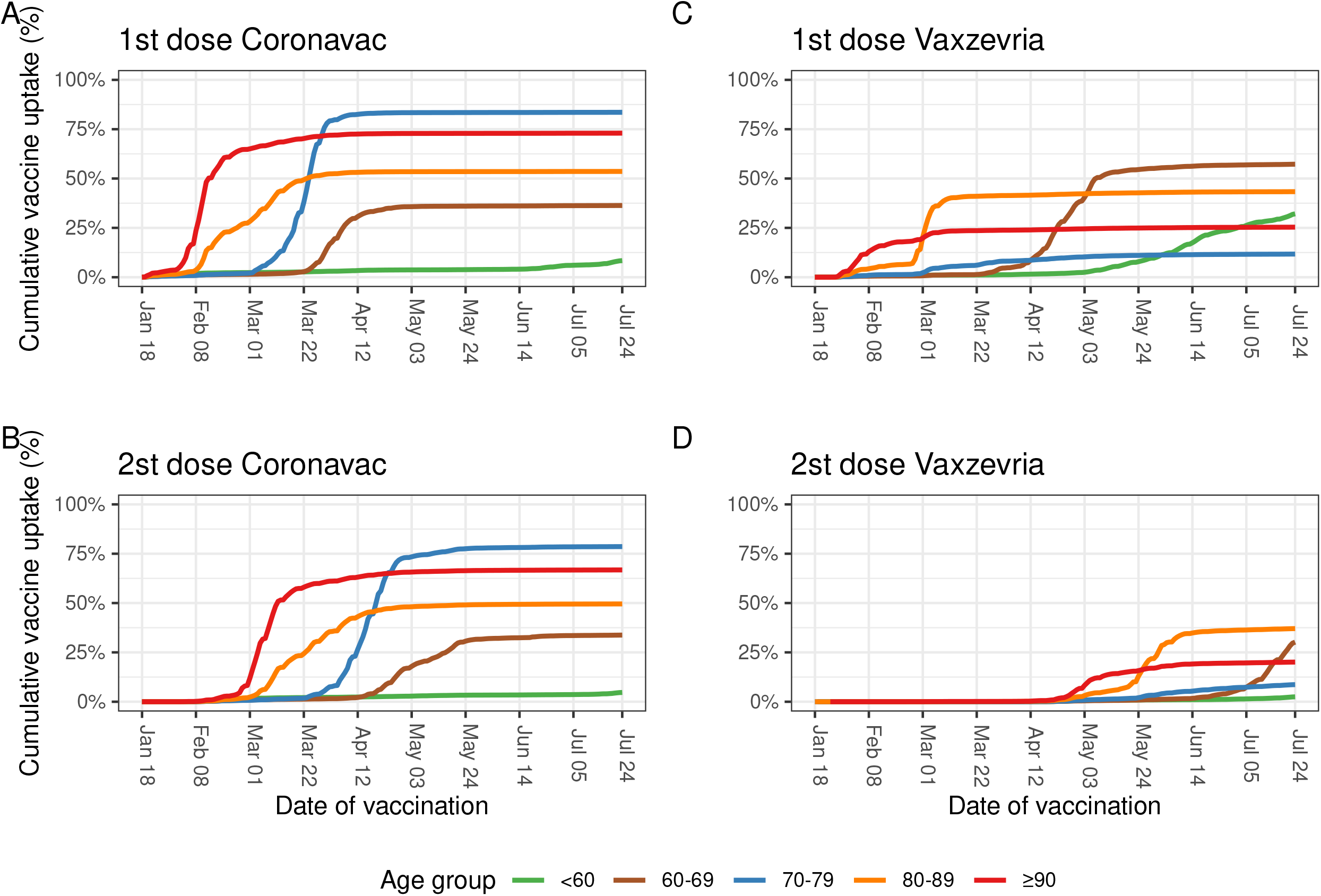
Coverage of first and second dose of CoronaVac and Vaxzevria in Brazil during the study period. The panels A, B, C and D shown the rate and coverage of the vaccination program regarding CoronaVac and Vaxzevria, A and C regarding first dose between January 18 and July 24 and panels B and D the second dose until July 24 2021.

Table 1 shows the COVID-19 VE analysis results, including number of events and incidence rate per 1,000 person-days and supplementary table S2 shows the crude and adjusted VE analysis. Individuals with full vaccination schedule (≥ 14 days after the second dose) with Vaxzevria had a 72.9% (95% CI 71.9 to 73.8) lower risk of infection, 88.0% (95% CI 86.8 to 89.2) lower risk of hospitalization, 89.1% (95% CI 87.0 to 90.8) lower risk of ICU admission, and 90.2% (95% CI 88.5 to 91.5) lower risk of death. Partial vaccination (i.e., ≥14 days after the first dose up to the second dose) with Vaxzeria was associated with a 34.7% lower risk of infection (95% CI 34.2 to 35.3) and at least 50% lower risk of hospitalization (55.2%; 95% CI 54.1 to 56.3), ICU admission (56.2.6%; 95% CI 54.4 to 58.0), and death (51.1%; 95% CI 49.0 to 53.0). Complete vaccination with CoronaVac was associated with lower risk of infection (52.7%, 95% CI 52.1-53.4), hospitalization (72.8%, 95% CI 71.8 to 73.7), ICU admission (73.8%, 95% CI 72.2 to 75.2) and death (73.7%, 95% CI 72.3 to 75.0). Partial vaccination with CoronaVac was associated with slight reduction in the risk of infection (18.6%; 95% CI 17.6 to 19.6), hospitalization (28.1%; 95% CI 26.3 to 29.9), ICU admission (28.5%; 95% CI 25.4 to 31.4), and death (29.4%; 95% CI 26.7 to 31.9).

**Table 1.**
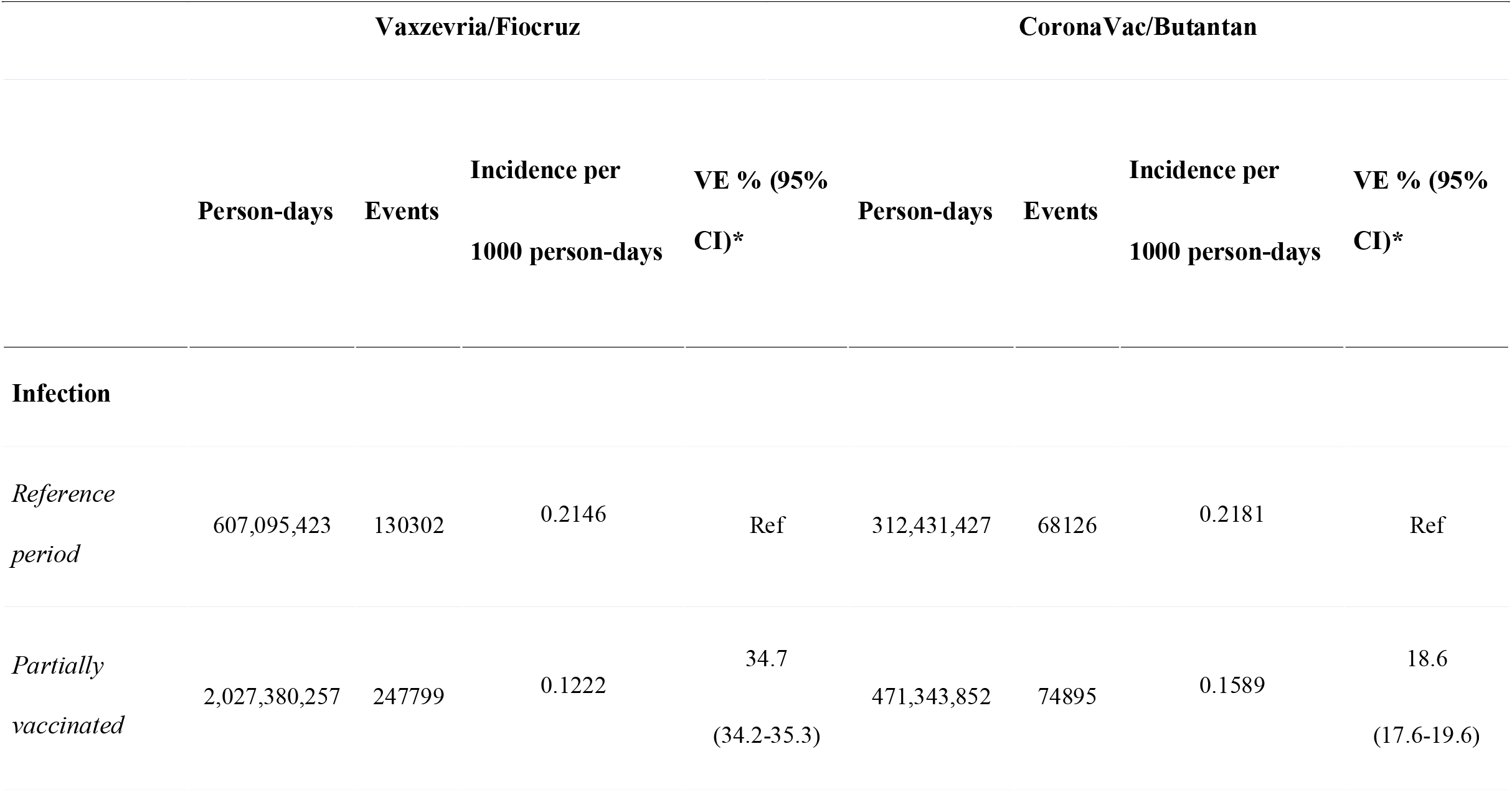
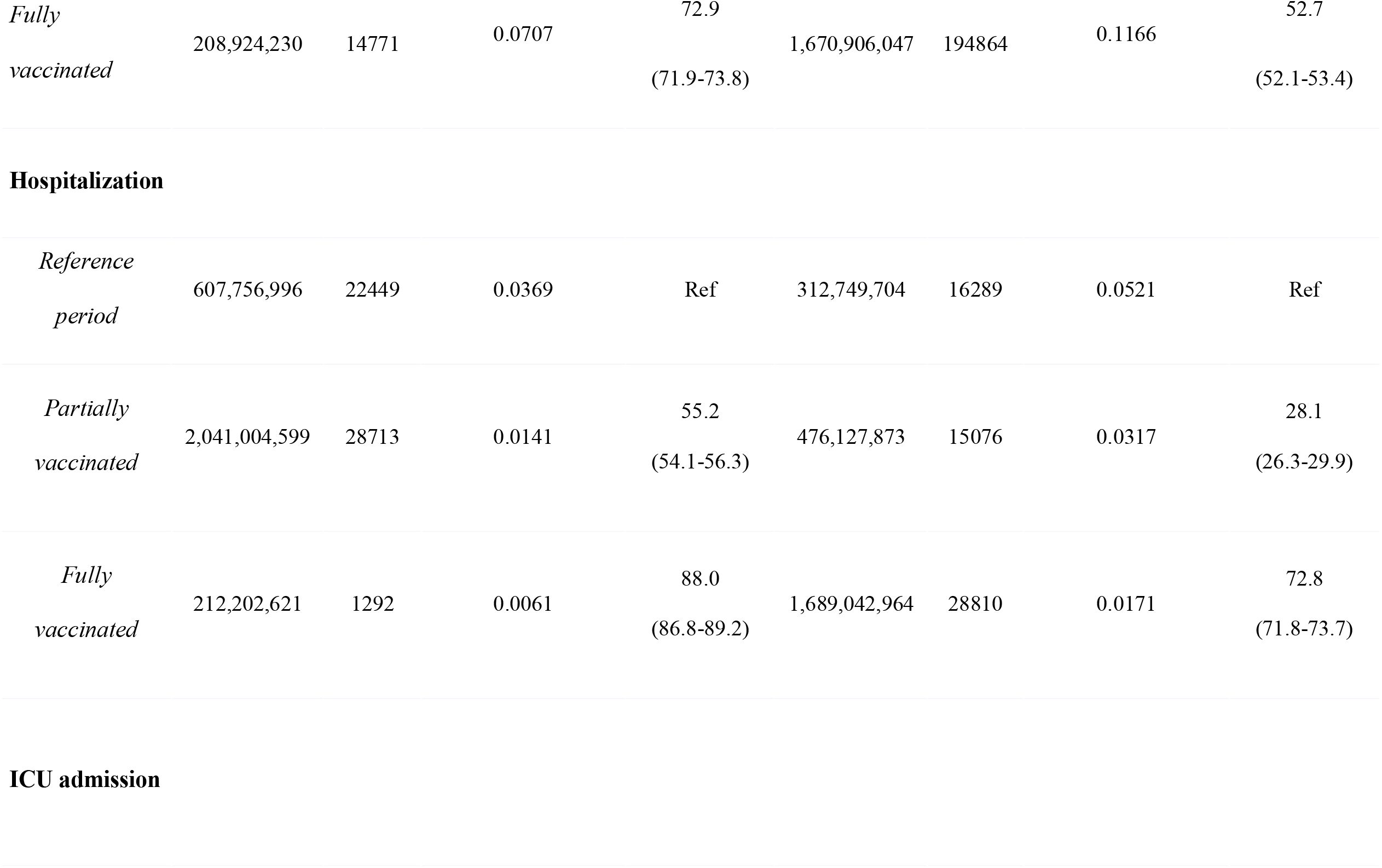
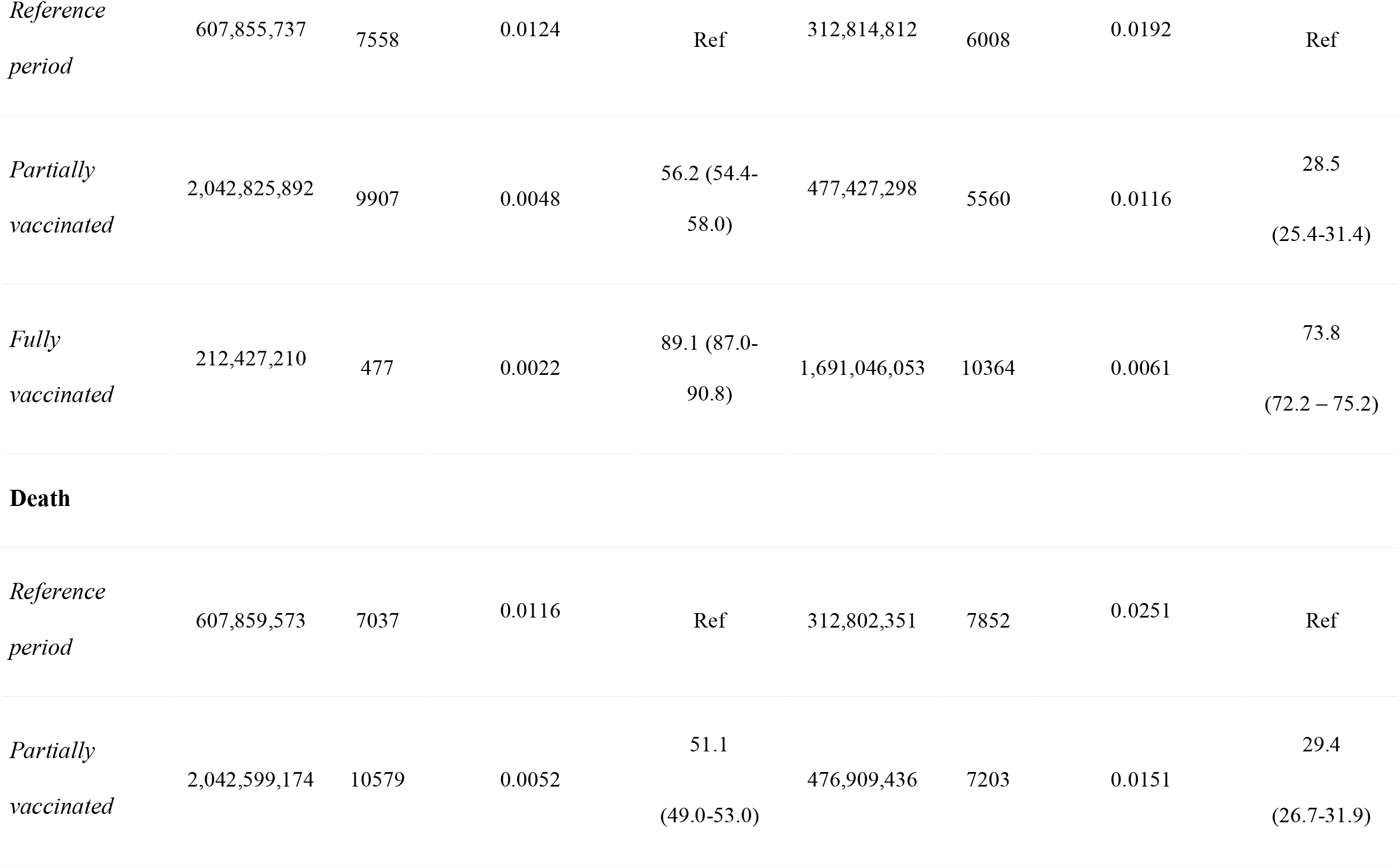
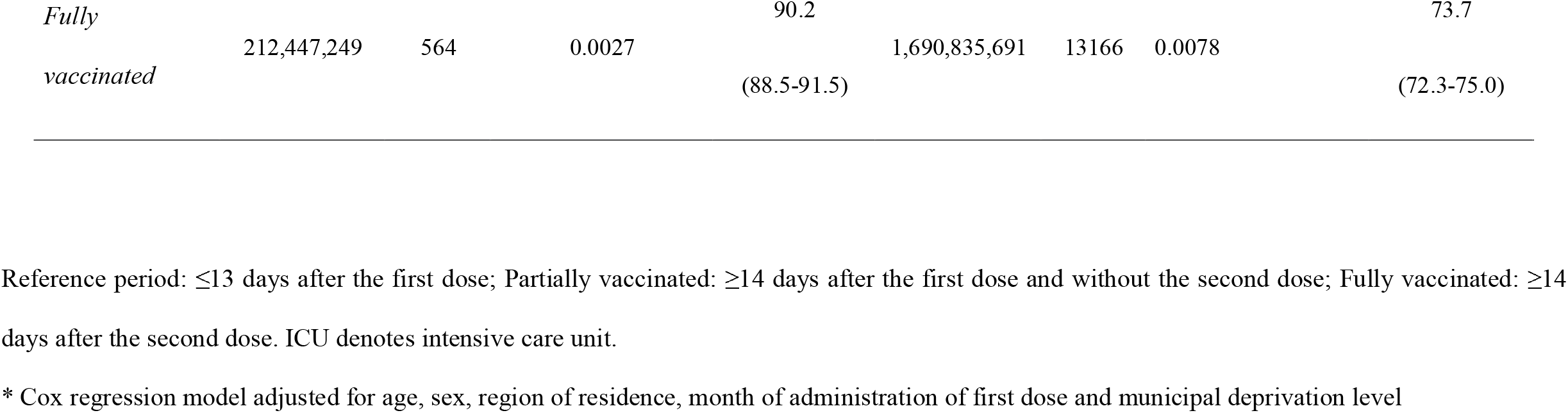
Vaccine effectiveness of Vaxzevria and CoronaVac in Brazil for COVID-19 infection, hospitalization, ICU admission, and death.

When stratifying the analysis by age, complete vaccination with Vaxzevria or CoronaVac allowed different degree of protection. Vaxzevria-induced VE was around 90% in distinct outcomes up to 89 years, whereas CoronaVac VE reached around 75% protection up to 79 years, with a modest decline in the 80-89 years. In individuals aged 90 years or older complete vaccination with Vaxzevria presented VE against death of 65.4%, whereas full vaccination with CoronaVac exhibited a VE of 33.6%, and only in this age group partial vaccination with CoronaVac exhibited no protection (Table S3, Figure 3).

**Figure 3.**
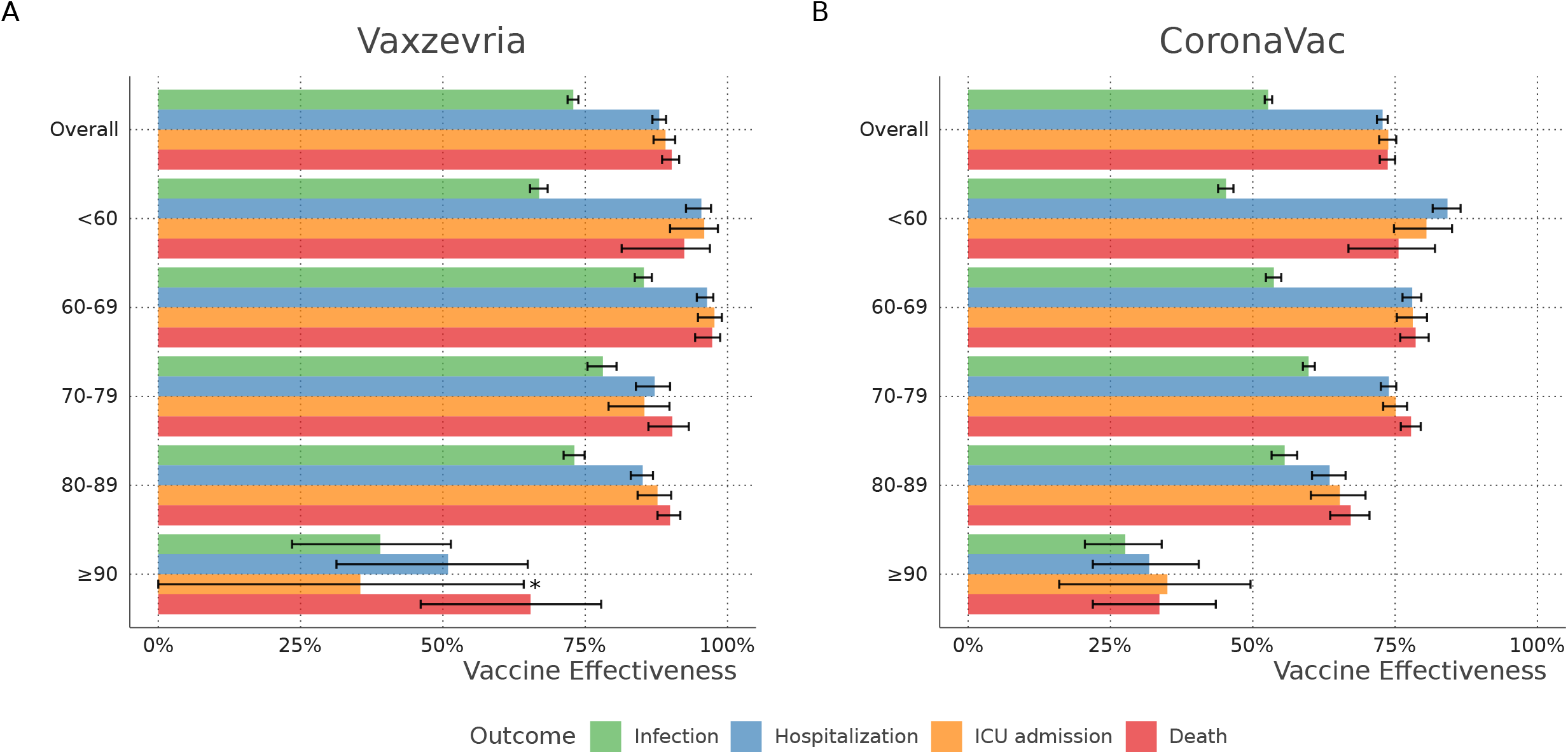
Vaccine effectiveness of Vaxzevria and CoronaVac in Brazil by age group. VE (1-Hazard Ratio) was obtained through Cox regression adjusted for age, sex, region of residence, the month of administration of first dose, and municipal deprivation level (IBP). *The point estimate and confidence interval for ICU admission in ≥90 y.o. are 35.5 (95%CI - 16.4 to 64.2%).

In order to further explore the impact of age on vaccine effectiveness and duration of protection, we analyzed the daily incidence of hospitalizations for the previously defined age groups for up to three months. CoronaVac vaccination was associated with decrease and maintenance of a low hospitalization incidence up to 84 days in vaccinees up to 79 years. The 80-89 and ≥90 age groups reached the lowest incidence 28 days after the 2nd dose. Incidence levels gradually increased afterward but were lower than those observed during the reference period or for partially vaccinated individuals (Figure 4A and Table S4).

**Figure 4.**
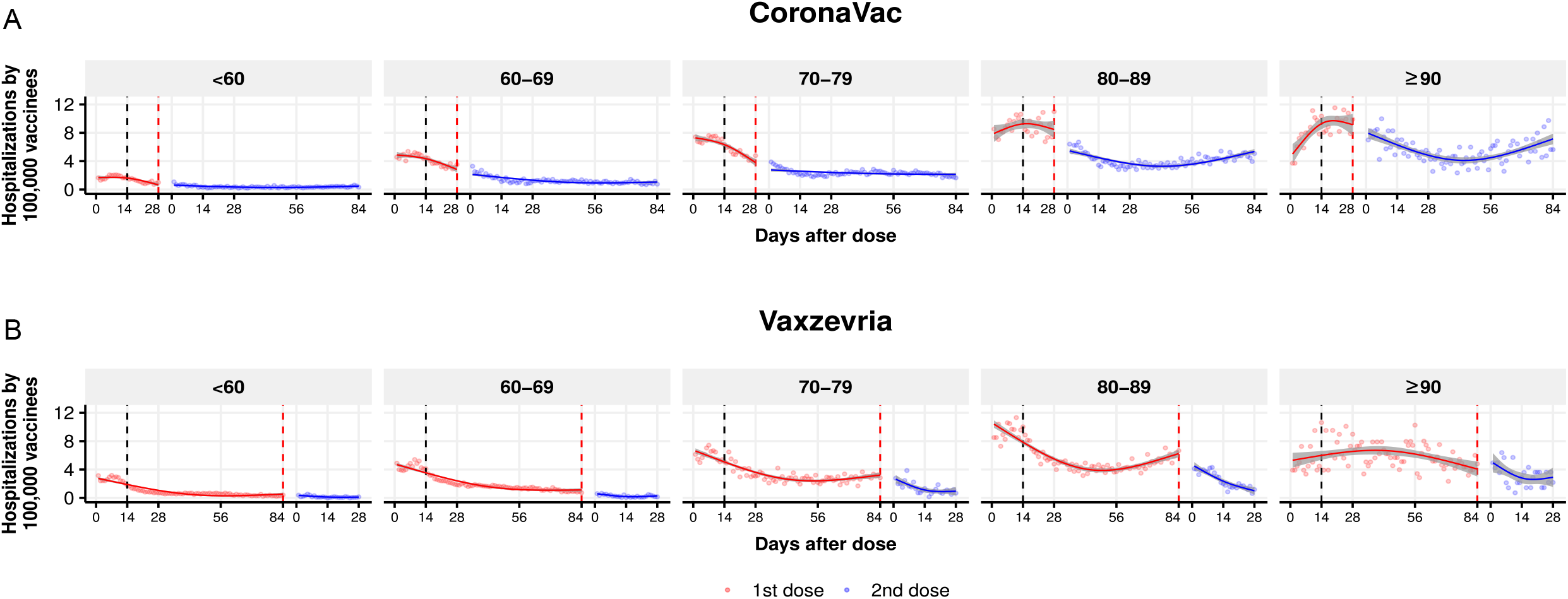
Incidence by day of new cases requiring hospitalization by 100,000 vaccinees, considering the difference between vaccination date and symptom on set. Dotted black line represent day 14, and dotted red line represent the ideal day of second dose (day 28 for CoronaVac and 84 for Vaxzevria). The lines represent restricted cubic spline with three degree of freedom, and the shaded areas 95% confidence intervals. Number at risk in each time point are presented in supplementary table S3. A) CoronaVac/Butantan vaccine, B) Vaxzevria/Fiocruz vaccine.

A single dose of Vaxzevria steadily reduced the incidence of hospitalization up to three months after the first dose, the period recommended for the administration of 2nd dose in individuals up to 79 years. In the age ranges of 80-89 years, there was an upward trend in incidence starting at day 56 after the 1st dose and before administering the second dose. Individuals ≥90 years maintained a constant incidence of hospitalization between 4 and 7/100.000 vaccinees after administering the first dose, that decreased after the second (Figure 4B and Table S4).

For both vaccines, there is an apparent rise, albeit at distinct levels, in the basal hospitalization incidence level in each life decade analyzed, highlighting the critical impact of age on the effectiveness of two vaccines that employ different technologies.

The comparison between the reference period of each vaccine shows no difference in the risk of infection, hospitalization, ICU admission, and death among the two vaccine groups (Table S5). Modifying the reference period for of up to 10 days after the first dose, we found VE point and interval estimates similar to those found in the primary analysis for both Vaxzeria and Coronavac vaccines (Table S6). Additionally, analysis of the outcomes clinical suspected cases in addition to laboratory confirmed cases were qualitatively equal to those found with only laboratory confirmed cases(Table S7).

## DISCUSSION

Analyzing data from almost 76 million vaccinated individuals, we demonstrate that following full vaccination Vaxzevria offered approximately 90.0% effectiveness against hospitalization, ICU admission, and death, while CoronaVac provided approximately 75% protection. Our findings regarding the Coronovac/Butantan vaccine protection against symptomatic COVID-19 are compatible with a previous Brazilian efficacy study^19,25^, but lower than reported by a Turkish efficacy trial.^8^ The effectiveness determined by a cohort study in Chile was higher than our findings for infection and hospitalization, but the differences may be partially explained by the higher frequency of younger individuals in the Chile study (51.2% vs. 38.5% of individuals younger than 60 years old). During the vaccination campaign, Brazil experienced a health system collapse, which may have influenced death rates.^26^ Additionally, the Gamma variant has been estimated at 28.6% in Chile and 69.6% in Brazil during the study periods.^1,12^ Plasma from fully vaccinated CoronaVac vaccinees exhibited a reduced capacity to neutralize the Gamma variant.^1^ Furthermore, vaccination in Chile was speedier than in Brazil what may have resulted in differences in viral transmission.^1^

For Vaxzevria, our findings of 72.9% effectiveness against infection exceeded the levels of 66.7% effectiveness reported in a combined analysis of four clinical trials conducted in the UK, South Africa, and Brazil.^7^ Effectiveness against hospitalization was consistent with the 80% and 88% protection observed in studies in Scotland^3^ and England,^11^ respectively. Additionally, our findings support the high level of protection offered by Vaxzevria despite the large circulation of the Gamma variant in Brazil during this period. Some studies have reported on the VE of Vaxzevria in populations infected by VOCs, mainly focused on protection against symptomatic infection or hospitalization.^9,10,13,20^ The findings reported herein, combined with data in the literature, confirm a consistently high rate of protection against moderate to severe COVID-19 in real-world studies, despite a large circulation of VOCs.

The protection conferred by the analyzed vaccines varied according to age group and declined in individuals with >80 years. It is evident a stepwise increase in the hospitalization incidence rate at each decade of life. Also noteworthy is the observed increase in the hospitalization incidence rate after approximately 60 days of administering the CoronaVac second dose in individuals >80 years, signaling for an early protective immunity waning. The short follow-up after the Vaxzevria second dose does not allow for estimating waning immunity with this vaccine. It is reasonable to attribute the observed reduction in effectiveness to immunosenescence, which is commonly associated with a higher frequency of comorbidities and may imply higher death rates. In the context of limited vaccine availability, the precise identification of age limits at which point immune protection becomes impaired can provide valuable evidence to inform public health measures. Considering the current scenario in Brazil, our findings demonstrate the eventual need for a vaccine booster dose in individuals >80 years who received CoronaVac and for individuals over 90 years immunized with Vaxzevria.

The differences in effectiveness between Vaxzevria and CoronaVac may be related to the different technologies used in these two products and how they influence immunogenicity.^27,28^

A relevant strength of our study is its large sample size, which allowed identifying the age limits in which immune protection becomes impaired. However, our study is also subject to some limitations. First, as VE was estimated using observational data, our analysis is subject to data availability and, therefore, to potential confounders. Although our analyses were not controlled for comorbidities, crude and adjusted VE estimates were similar. Additionally, comorbidities are in the causal pathway between age and COVID-19 severity. Therefore, by controlling for age, we are also indirectly controlling for comorbidities.^29^ Second, in contrast to many VE studies, the reference period used herein for comparison purposes was 1-13 days after vaccination. Although using early post-vaccination as a reference may underestimate VE, previous studies have used a similar approach and obtained results similar to those found in clinical trials.^30,31^ The early post-vaccination period can also be used as a bias indicator related to differences in SARS-CoV-2 infection risk. Additionally, the effectiveness results of the present report are similar, in the pertinent age ranges, to reports on both vaccines using distinct approaches.^1,19,20^ Finally, we also performed a sensitivity analysis, which demonstrated similar results when a 0–10-day reference period was applied.

Using the data available in Brazil, we estimated the overall VE for each vaccine evaluated and by age group. Vaxzevria/Fiocruz and CoronaVac/Butantan were both shown to be highly protective against severe COVID-19 in the population aged up to 80 years. Due to decreased VE, an early booster dose may be considered for those over 80 years of age who received CoronaVac, especially for individuals over 90 years, regardless of which of these two vaccines were administered. Despite high population adherence, the vaccination campaign is evolving unevenly throughout Brazil, and continuous monitoring of VE in the current context may provide sound evidence to inform public health measures.

## Data Availability

We used third-party data, provided by the Brazilian Ministry of Health. Any request for access to the data shall be directed to DATASUS - Ministry of Health Brazil:
https://datasus.saude.gov.br/

https://datasus.saude.gov.br/

## ETHICAL CONSIDERATIONS

The Brazilian National Commission in Research Ethics approved the research protocol (CONEP approval number 4.921.308). All work presented here used unidentified secondary data in accordance with the Brazilian Personal Data Protection General Law (LGPD). Data was manipulated in a secure computing environment, ensuring protection against data leakage and records re-identification.

## DATA SHARING

We used third-party data, provided by the Brazilian Ministry of Health. Any request for access to the data shall be directed to DATASUS - Ministry of Health Brazil: https://datasus.saude.gov.br/

## ACKNOWLEDGMENTS

The authors thank DATASUS for its excellent work in providing unidentified databases. GW, MB, and MB-N are research fellows from CNPq, the Brazilian National Research Council. The authors would also like to thank Andris K. Walter for English language revision and manuscript copy-editing assistance.

