## Supplementary for "Influence of age on the effectiveness and duration of protection in Vaxzevria and CoronaVac vaccines"

Table S1. Demographic characteristics of individuals that received at the first dose of Vaxzevria and CoronaVac in Brazil between January 18 and July 24, 2021.

|  | Vaxzevria/Fiocruz | | | CoronaVac/Butantan | | | | |
| --- | --- | --- | --- | --- | --- | --- | --- | --- |
|  | Persons with only one dose N=40,047,771  n (%) | Persons with two doses N=10,120,116  n (%) | Total  N=50,167,827  n (%) | Persons with only one dose N=5,411,455  n (%) | Persons with two doses N=20,340,558  n (%) | | Total  N=25,752,013  n (%) | |
| Sex (Female) | 20,982,634 (52.4) | 5,832,823 (57.6) | 26,815,457 (53.5) | 2,818,107 (52.1) | 12,020,835 (59.1) | | 14,838,942(57.6) | |
| Age group |  |  |  |  |  | |  | |
| *<20* | 391,372 (1.0) | 38,671 (0.4) | 430,043 (0.9) | 50,236 (0.9) | 79,260 (0.4) | | 129,496 (0.4) | |
| *20-29* | 3,117,341 (7.8) | 473,392 (4.7) | 3,590,733 (7.2) | 506,589 (9.4) | 1,021,888 (5.0) | | 1,528,477 (5.9) | |
| *30-39* | 7,410,578 (18.5) | 706,387 (7.0) | 8,116,965 (16.2) | 2,239,054 (41.4) | 1,420,631 (7.0) | | 3,659,685 (14.2) | |
| *40-49* | 11,090,699 (27.7) | 771,016 (7.6) | 11,861,715 (23.6) | 1,091,293 (20.2) | 1,703,783 (8.4) | | 2,795,076 (10.9) | |
| *50-59* | 13,080,834 (32.7) | 947,686 (9.4) | 14,028,520 (28.0) | 465,405 (8.6) | 1,343,148 (6.6) | | 1,808,553 (7.0) | |
| *60-69* | 4,428,849 (11.1) | 4,962,583 (49.0) | 9,391,432 (18.7) | 426,909 (7.9) | 5,556,410 (27.3) | | 5,983,319 (23.2) | |
| *70-79* | 266,237 (0.7) | 761,801 (7.5) | 1,028,038 (2.0) | 443,235 (8.2) | 6,991,138 (34.4) | | 7,434,373 (28.9) | |
| *80-89* | 224,223 (0.6) | 1,316,632 (13.0) | 1,540,855 (3.1) | 144,512 (2.7) | 1,752,396 (8.6) | | 1,896,908 (7.4) | |
| *≥90* | 37,578 (0.1) | 141,948 (1.4) | 179,526 (0.4) | 44,222 (0.8) | 471,904 (2.3) | | 516,126 (2.0) | |
| Region of residence |  |  |  |  |  | |  | |
| *Central West* | 2,879,201 (7.2) | 834,792 (8.2) | 3,713,993 (7.4) | 283,168 (5.2) | | 1,468,272 (7.2) | | 1,751,440 (6.8) |
| *Northeast* | 9,623,648 (24.0) | 2,384,870 (23.6) | 12,008,518 (23.9) | 1,014,849 (18.8) | | 4,696,346 (23.1) | | 5,711,195 (22.2) |
| *North* | 3,030,135 (7.6) | 811,155 (8.0) | 3,841,290 (7.7) | 313,351 (5.8) | | 1,242,076 (6.1) | | 1,555,427 (6.0) |
| *Southeast* | 18,058,452 (45.1) | 4,179,631 (41.3) | 22,238,083 (44.3) | 3,254,684 (60.1) | | 9,585,931 (47.1) | | 12,840,615 (49.9) |
| *South* | 6,246,820 (15.6) | 1,853,770 (18.3) | 8,100,590 (16.1) | 512,943 (9.5) | | 3,234,932 (15.9) | | 3,747,875 (14.6) |
| *Missing* | 209,455 (0.5) | 55,898 (0.6) | 265,353 (0.5) | 32,460 (0.6) | | 113,001 (0.6) | | 145,461 (0.6) |
| Brazilian Municipal Deprivation Index |  |  |  |  | |  | |  |
| *1* | 7,919,430 (19.8) | 2,288,455 (22.6) | 10,207,885 (20.3) | 1,230,281 (22.7) | | 4,499,885 (22.1) | | 5,730,166 (22.3) |
| *2* | 7,720,017 (19.3) | 1,858,486 (18.4) | 9,578,503 (19.1) | 1,406,660 (26.0) | | 4,221,255 (20.8) | | 5,627,915 (21.9) |
| *3* | 8.277,771 (20.7) | 2,050,852 (20.3) | 10,328,623 (20.6) | 1,067,535 (19.7) | | 4,147,004 (20.4) | | 5,214,539 (20.2) |
| *4* | 8,224,392 (20.5) | 1,922,733 (19.0) | 10,147,125 (20.2) | 782,115 (14.5) | | 3,851,004 (18.9) | | 4,633,119 (18.0) |
| *5* | 7,696,646 (19.2) | 1,943,692 (19.2) | 9,640,338 (19.2) | 892,404 (16.5) | | 3,508,409 (17.2) | | 4,400,813 (17.1) |
| *Missing* | 209,455 (0.5) | 55,898 (0.6) | 265,353 (0.5) | 32,460 (0.6) | | 113,001 (0.6) | | 145,461 (0.6) |

The study participants were included if they received first dose of CoronaVac of Vaxzevria between January 18 and July 24, 2021. The Brazilian Municipal Deprivation Index works as proxy for socioeconomic status.

Table S2. Crude and adjusted Vaccine effectiveness of Vaxzevria and CoronaVac in Brazil for COVID-19 infection, hospitalization, ICU admission and death.

|  | Vaxzevria/Fiocruz | | CoronaVac/Butantan | |
| --- | --- | --- | --- | --- |
|  | CRUDE VE %  (95% CI) | ADJUSTED VE %  (95% CI)* | CRUDE VE %  (95% CI) | ADJUSTED VE %  (95% CI)* |
| Infection |  |  |  |  |
| *Reference period* | — | — | — | — |
| *Partially vaccinated* | 30.9 (30.3-31.5) | 34.7 (34.2-35.3) | 15.9 (14.9-16.9) | 18.6 (17.6-19.6) |
| *Fully vaccinated until 13 days* | 64.3 (63.5-65.1) | 65.5 (64.6-66.3) | 36.5 (35.7-37.3) | 40.1 (39.3-40.9) |
| *Fully vaccinated* | 70.5 (69.6-71.4) | 72.9 (71.9-73.8) | 50.0 (49.3-50.7) | 52.7 (52.1-53.4) |
| Hospitalization |  |  |  |  |
| *Reference period* | — | — | — | — |
| *Partially vaccinated* | 51.7 (50.6-52.8) | 55.2 (54.1-56.3) | 25.1 (23.2-26.9) | 28.1 (26.3-29.9) |
| *Fully vaccinated until 13 days* | 71.5 (69.7-73.2) | 79.9 (78.6-81.2) | 52.7 (51.3-54.1) | 57.6 (56.4-58.9) |
| *Fully vaccinated* | 81.9 (80.1-83.6) | 88.0 (86.8-89.2) | 69.2 (68.2-70.3) | 72.8 (71.8-73.7) |
| ICU admission |  |  |  |  |
| *Reference period* | — | — | — | — |
| *Partially vaccinated* | 52.6 (50.6-54.5) | 56.2 (54.4-58.0) | 25.2 (22.1-28.3) | 28.5 (25.4-31.4) |
| *Fully vaccinated until 13 days* | 70.0 (66.8-73.0) | 80.3 (78.1-82.3) | 53.0 (50.7-55.2) | 58.2 (56.2–60.2) |
| *Fully vaccinated* | 82.4 (79.3-85.1) | 89.1 (87.0-90.8) | 70.0 (68.2-71.7) | 73.8 (72.2–75.2) |
| Death |  |  |  |  |
| *Reference period* | — | — | — | — |
| *Partially vaccinated* | 46.7 (44.4-48.8) | 51.1 (49.0-53.0) | 26.6 (23.9-29.2) | 29.4 (26.7-31.9) |
| *Fully vaccinated until 13 days* | 57.2 (53.2-60.8) | 79.4 (77.4-81.3) | 52.6 (50.6-54.6) | 58.6 (56.8-60.3) |
| *Fully vaccinated* | 75.6 (71.7-78.9) | 90.2 (88.5-91.5) | 69.1 (67.5-70.6) | 73.7 (72.3-75.0) |
| * Cox regression model adjusted for age, sex, region of residence, month of administration of first dose and municipal deprivation level. | | | | |

**Table S3. Vaccine effectiveness of Vaxzevria and CoronaVac in Brazil by age groups for COVID-19 infection, hospitalization, ICU admission and death.**

|  | | **Vaxzevria/Fiocruz** | | | | | **CoronaVac/Butantan** | | | | |
| --- | --- | --- | --- | --- | --- | --- | --- | --- | --- | --- | --- |
|  | | <60 | 60-69 | 70-79 | 80-89 | ≥90 | <60 | 60-69 | 70-79 | 80-89 | ≥90 |
| Infection | |  |  |  |  |  |  |  |  |  |  |
|  | *Partially*  *vaccinated* | 39.9  (39.3-40.5) | 17.9  (16.3-19.4) | 24.7  (19.8-29.3) | 25.6  (22.2-28.9) | -58.1  (-86.3- -34.1) | 22.1  (20.5-23.6) | 14.3  (12.4-16.4) | 23.8  (22.1-25.5) | 1.1  (-3.1 --5.1) | -23.7  (-34.5- -13.8) |
| *Fully vaccinated* | | 66.9  (65.3-68.4) | 85.3  (83.7-86.7) | 78.1  (75.4-80.5) | 73.1  (71.2-74.9) | 39.0  (23.5-51.4) | 45.3  (43.9-46.6) | 53.7  (52.3-55.0) | 59.8  (58.8-60.9) | 55.6  (53.3-57.8) | 27.6  (20.5-34.0) |
| Hospitalization | |  |  |  |  |  |  |  |  |  |  |
| *Partially vaccinated* | | 65.7  (64.5-66.9) | 44.8  (42.3-47.1) | 33.0  (25.5-39.7) | 32.6  (27.8-37.1) | -34.3  (-69.6- -6.3 | 44.7  (40.2-48.8) | 29.3  (25.7-32.8) | 32.3  (29.7-34.9) | 8  (2.0-13.6) | -17.6  (-32.8- -4.2) |
|  | *Fully vaccinated* | 95.4  (92.7-97.1) | 96.4  (94.6-97.5) | 87.2  (83.9-89.9) | 85.1  (83.0-86.9) | 50.9  (31.3-64.9) | 84.2  (81.6-86.5) | 78.0  (76.3-79.6) | 73.9  (72.5-75.2) | 63.5  (60.4-66.3) | 31.8  (21.9-40.5) |
| ICU admission | |  |  |  |  |  |  |  |  |  |  |
| *Partially vaccinated* | | 66.1  (64.0-68.1) | 48.0  (44.0-51.6) | 38.5  (26.7-48.4) | 34.7  (26.2-42.0) | -50.1  (-131.6 -2.8) | 43.0  (34.4-50.6) | 28.1  (22.2-33.6) | 31.8  (27.4-35.8) | 18.6  (9.3-27.0) | -29.9  (-61.9- -4.2) |
| *Fully vaccinated* | | 95.9  (89.9-98.3) | 97.7  (94.8 – 99.0) | 85.4  (79.1 – 89.8) | 87.5  (84.2-90.1) | 35.5  (-16.4 - 64.2) | 80.5  (74.8-85.0) | 78.1  (75.3-80.6) | 75.1  (72.9-77.1) | 65.3  (60.2-69.8) | 35.0  (16.0-49.6) |
|  | Death |  |  |  |  |  |  |  |  |  |  |
| *Partially vaccinated* | | 65.7  (63.3-68.1) | 44.8  (40.6-48.6) | 36.7  (26.8-45.2) | 37.8  (32.0-43.1) | -44.1  (-88.0- 10.3) | 50.7  (39.7-59.7) | 34.9  (29.5-39.8) | 37.7  (34.2-40.9) | 9.6  (2.2-16.4) | -23.7  (-42.5- -7.4) |
| *Fully vaccinated* | | 92.4  (81.4-96.9) | 97.3  (94.3-98.7) | 90.3  (86.1-93.2) | 89.9  (87.7-91.7) | 65.4  (46.1 – 77.8) | 75.6  (66.8-82.0) | 78.6  (75.9-80.9) | 77.8  (76.1-79.5) | 67.2  (63.6-70.5) | 33.6  (21.9-43.5) |

*Obtained through Cox regression model adjusted for age, sex, region of residence, month of administration of first dose and municipal deprivation level

Table S4. Number at risk in each time point and age group of Vaxzevria and CoronaVac vaccinees.

| **Vaccine/Dose** | | **Number at risk -n(%)** | |  | |
| --- | --- | --- | --- | --- | --- |
| **Vaxzevria 1st dose** | 0 | 14 | 28 | 56 | 84 |
| <60 | 38027976 | 33148016  (87.2) | 27806653  (73.1) | 11310228  (29.7) | 2236198  (5.9) |
| 60-69 | 9391432 | 9348405  (99.5) | 9298903  (99.0) | 8980737  (95.6) | 5224876  (55.6) |
| 70-79 | 1028038 | 1019946  (99.3) | 1010596  (98.3) | 966268  (94.0) | 740381  (72.1) |
| 80-89 | 1540855 | 1535589  (99.8) | 1531174  (99.5) | 1511950  (98.2) | 1304433  (84.7) |
| ≥90 | 179526 | 178820  (99.6) | 178124  (99.2) | 175464  (97.7) | 145978  (81.3) |
| **Vaxzevria 2nd dose** | 0 | 14 | 28 |  |  |
| <60 | 2935556 | 1895514  (64.6) | 1406733  (47.9) | — | — |
| 60-69 | 4957980 | 1845510  (37.2) | 647419  (13.1) | — | — |
| 70-79 | 760757 | 682485  (89.7) | 594316  (78.1) | — | — |
| 80-89 | 1313953 | 1296936  (98.7) | 1277385  (97.2) | — | — |
| ≥90 | 141713 | 139609  (98.5) | 137651  (97.1) | — | — |
| **Coronavac 1st dose** | 0 | 14 | 28 |  |  |
| <60 | 9921287 | 7427329  (74.9) | 3125644  (31.5) | — | — |
| 60-69 | 5983319 | 5962455  (99.8) | 3656668  (61.2) | — | — |
| 70-79 | 7434373 | 7409167  (99.9) | 3290219  (44.4) | — | — |
| 80-89 | 1896908 | 1885159  (99.8) | 846137  (44.8) | — | — |
| ≥90 | 516126 | 512039  (99.9) | 222304  (43.4) | — | — |
| **Coronavac 2nd dose** | 0 | 14 | 28 | 56 | 84 |
| <60 | 5567184 | 4412190  (79.3) | 4215556  (75.7) | 3998650  (71.8) | 3341667  (60.0) |
| 60-69 | 5552219 | 5525959  (99.5) | 5486636  (98.8) | 5217451  (94.0) | 2788211  (50.2) |
| 70-79 | 6985451 | 6976252  (99.9) | 6964850  (99.7) | 6914090  (99.0) | 6496505  (93.0) |
| 80-89 | 1750741 | 1748011  (99.8) | 1745387  (99.7) | 1736204  (99.2) | 1696755  (96.9) |
| ≥90 | 471521 | 470763  (99.8) | 470092  (99.7) | 468313  (99.3) | 461983  (98.0) |

Table S5. Comparison between reference groups of each vaccine.

| **Outcome** | **Hazard Ratio* (95% CI)** |
| --- | --- |
| **Infection** |  |
| Vaxzevria/Fiocruz | ─ |
| CoronaVac/Butantan | 1.005 (0.993 - 1.018) |
| **Hospitalization** |  |
| Vaxzevria/Fiocruz | ─ |
| CoronaVac/Butantan | 0.987 (0.961 - 1.015) |
| **ICU admission** |  |
| Vaxzevria/Fiocruz | ─ |
| CoronaVac/Butantan | 1.023 (0.977-1.071) |
| **Death** |  |
| Vaxzevria/Fiocruz | ─ |
| CoronaVac/Butantan | 1.000 (0.958-1.042) |

Cox regression model adjusted for age, sex, region of residence, month of administration of first dose and municipal deprivation level.

Table S6. Robustness analysis with different time windows as reference period

|  | Vaxzevria/Fiocruz VE % (95% CI) | CoronaVac/Butantan VE % (95% CI) |
| --- | --- | --- |
| Reference Period: | 0-10 days | 0-10 days |
| Infection |  |  |
| *Partially vaccinated* | 34.9 (34.2-35.6) | 18.0 (16.8-19.3) |
| *Fully vaccinated until 13 days* | 67.2 (66.2-68.2) | 38.8 (37.8-39.8) |
| *Fully vaccinated* | 74.4 (73.3-75.5) | 54.2 (53.3-55.0) |
| Hospitalization |  |  |
| *Partially vaccinated* | 54.2 (53.0-55.4) | 27.0 (24.9-29.0) |
| *Fully vaccinated until 13 days* | 79.1 (77.7-80.5) | 55.9 (54.4-57.3) |
| *Fully vaccinated* | 88.3 (86.9-89.5) | 72.7 (71.6-73.7) |
| ICU admission |  |  |
| *Partially vaccinated* | 55.8 (53.8-57.8) | 28.0 (24.7-31.2) |
| *Fully vaccinated until 13 days* | 79.6 (77.2-81.8) | 56.9 (54.5-59.2) |
| *Fully vaccinated* | 89.5 (87.3-91.4) | 73.8 (72.0-75.4) |
| Death |  |  |
| *Partially vaccinated* | 50.1 (47.8-52.3) | 28.6 (25.7-31.4) |
| *Fully vaccinated until 13 days* | 78.6 (76.4-80.7) | 57.8 (55.7-59.7) |
| *Fully vaccinated* | 90.0 (88.2-91.5) | 73.5 (71.9-74.9) |

Cox regression model adjusted for age, sex, region of residence, month of administration of first dose and municipal deprivation level.

Table S7: Percentage of events with laboratory confirmation and VE using all cases (laboratory and clinical suspected)

| **Vaxzevria/Fiocruz** | | | | | | **Coronavac/Butantan** | | | | |
| --- | --- | --- | --- | --- | --- | --- | --- | --- | --- | --- |
|  | **Events- Laboratory Confirmed** | Events-Confirmed or Clinical Suspected | % Confirmed | | VE*  (95% CI) | | **Events- Laboratory Confirmed** | Events-Confirmed or Clinical Suspected | % Confirmed | VE*  (95% CI) |
| **Hospitalization** |  |  | |  |  | |  |  |  |  |
| *Reference period* | 22449 | 28394 | | 79.1 |  | | 16289 | 21133 | 77.1 |  |
| *Partially vaccinated* | 28713 | 38202 | | 75.2 | 53.5  (52.5 - 54.5) | | 15076 | 19975 | 75.5 | 26.9  (25.3 -28.5) |
| *Fully vaccinated* | 1292 | 1948 | | 66.3 | 86.6  (85.5 - 87.7) | | 28810 | 38180 | 75.5 | 71.5  (70.6 -72.4) |
| **ICU admission** |  |  | |  |  | |  |  |  |  |
| *Reference period* | 7558 | 9223 | | 81.2 |  | | 6008 | 7587 | 79.2 |  |
| *Partially vaccinated* | 9907 | 12742 | | 77.8 | 54.4  (52.7-56.1) | | 5560 | 7240 | 76.8 | 27.0  (24.3-29.6) |
| *Fully vaccinated* | 477 | 684 | | 69.7 | 87.8  (85.9-89.5) | | 10364 | 13345 | 77.7 | 72.7  (71.3 -74.1) |
| **Death** |  |  | |  |  | |  |  |  |  |
| Reference period | 7037 | 8733 | | 80.6 |  | | 7852 | 9974 | 78.7 |  |
| Partially vaccinated | 10579 | 13775 | | 76.8 | 49.5  (47.6-51.3) | | 7203 | 9293 | 77.5 | 28.7  (26.3-30.9) |
| Fully vaccinated | 564 | 807 | | 69.9 | 89.1  (87.6-90.4) | | 13166 | 16826 | 78,2 | 73.1  (71.9-74.2) |

Obtained through Cox regression model adjusted for age, sex, region of residence, month of administration of first dose and municipal deprivation level
